# Mendelian randomization suggests a causal link between glycemic traits and thoracic aortic structures and diseases

**DOI:** 10.1101/2024.09.18.24313344

**Authors:** Tselmen Daria, Kruthika Iyer, Hasan Alkhairo, Pik Fang Kho, Ken Suzuki, Konstantinos Hatzikotoulas, Lorraine Southam, Henry J Taylor, Xianyong Yin, Ravi Mandla, Alicia Huerta-Chagoya, Nigel W Rayner, Michael G. Levin, Scott M Damrauer, Philip S Tsao, James R Priest, James Pirruccello, Justin B Echouffo Tcheugui, Catherine Tcheandjieu

## Abstract

**Importance:** Type 2 diabetes mellitus (T2DM) and elevated glucose levels have been inversely associated with aortic aneurysms. However, the causality of this relationship remains uncertain. Additionally, there is a lack of studies investigating the association between glycemic traits and imaging-based thoracic aortic phenotypes.

**Objective:** We investigated whether T2DM and glycemic measures (fasting glucose (FG), fasting insulin (FI), glycated hemoglobin (HbA1c), and 2-hour post-load glucose (2hPG)) are causally associated with various imaging and clinical thoracic aortic phenotypes.

**Design, setting, and participants:** We performed Mendelian randomization (MR) analysis using summary statistics from genome-wide association studies of glycemic traits (GT) and aortic phenotypes. We employed a two-sample univariate MR (UVMR) followed by a multivariable MR and MR analysis using a cluster-based approach.

**Main Outcomes and Measures:** The outcome includes imaging-based ascending and descending aortic diameters (AAoD and DAoD), aortic distensibility (AoDist) and strain (AoStr), and thoracic aortic aneurysm and dissection (TAAD). The GT include FI, FG, HbA1c, 2hPG and T2DM.

**Results:** We observed an inverse association between 2hPG, FG, HbA1c, and T2DM and AAoD, DAoD. For instance, the genetically predicted increases levels of 2hPG, FG, HbA1c, and T2DM were associated with decreased AAoD (2hPG β= -0.2, p=3x10^-08^; FG β= -0.21, p=5x10^-05^; HbA1c β= -0.36, p=2x10^-07^; T2DM β=-0.04, p=2x10^-04^) and reduce risk TAAD (2hPG decreased TAAD (2hPG OR= 0.70, p=4x10^-04^; FG OR= 0.58, p=3x10^-05^; HbA1c OR= 0.62, p=5x10^-03^; T2DM OR= 0.90, p=6x10^-6^). Further investigation showed that the inverse association between T2DM and aortic phenotypes is driven by genetic predictors of T2DM beta cell proinsulin clusters. UVMR and Proteomic MR showed a strong association with aortic phenotypes for genes, such *as AGER, GLRX, TCF7L2*, and *GCK*, which are known to play an important role in glucose regulation.

**Conclusion and relevance:** Our findings suggest a potentially causal impact of GT on the aortic vasculature. Furthermore, specific glucose regulation genes such as *GCK*, *AGER*, and *TCF7L2* appear to contribute to this process, opening avenues for potentially leveraging the druggability of these genes for treatment or preventions of TAAD.

## INTRODUCTION

Aortic aneurysm and dissection involve weakening or balloon-like dilation in the aorta’s wall, often asymptomatic until dissection occurs. These dissections, associated with high mortality rates, result in death in over 90% of cases within 48 hours if untreated^1^. The only curative treatment available is surgical; drug-based preventive therapies are lacking, highlighting a huge unmet need for clinical practice. Thoracic aortic aneurysm and dissection (TAAD) is generally linked to genetic disorders of the extracellular matrix and the contractile apparatus but also shares cardiovascular risk factors including male gender, age, smoking, hypertension, and hyperlipidemia^2^. Deep learning has enabled large-scale cardiac phenotyping including aortic measures in biobanks like UK Biobank^3^. Conducting genome-wide association studies (GWAS) on Magnetic Resonance Imaging (MRI)-derived aortic measures has led to the identification of genetic markers linked to aortic morphology and function. These discoveries have enhanced our understanding of the biology and genetics of aortic aneurysms and dissections^4,5^. Intriguingly, epidemiological studies have shown an inverse association between type 2 diabetes (T2DM) and TAAD^6,7^, however, the causal nature of this relationship remained unknown. A better understanding of the mechanisms underlying the negative association could help the development of innovative diagnostic and therapeutic approaches.

Observational studies can be marred with various types of errors such as confounding, reverse causation, mediation bias, or inability to fully differentiate the direct effect of T2DM on aortic aneurysms independent of upstream factors such as obesity and hypertension. Moreover, extant epidemiological studies have seldom examined the full spectrum of GT related to T2DM, such as insulin markers, glycated hemoglobin (HbA1c), fasting blood glucose (FG), and 2hour post glucose load (2hPG), which may offer unique insight into the link between dysglycemia and aortic structure and TAAD. However, very few studies have examined imaging-based subclinical aortic structure, that could indicate earlier stages of the aortic disease process.

Mendelian randomization (MR) utilizes genetic variations associated with exposures to assess potentially causal relationships between exposures and outcomes while controlling for confounding factors^8^. We aimed to investigate whether T2DM and various glycemic traits (FG, FI, HbA1c, and 2hPG) affect thoracic aortic structure and subsequently influence the development of TAAD using a comprehensive multi-omic approach. This approach includes an MR analysis across multiple populations, an innovative clustering-based MR analysis, proteomic MR, transcriptome-wide association (TWAS), and gene/pathway enrichment analysis.

## METHODS

### Study population

All epidemiological analyses were conducted using data from the UK Biobank (UKBB) under application number 87,255. We analyzed approximately 30,000 participants who had undergone cardiac MRI, from which aortic phenotypes were extracted. To examine the association with TAAD, we utilized the full cohort of UKBB participants (1,135 cases /405,574 controls).

For the MR analysis, genetic summary statistics (SumStat) of associations between genetic variants and MRI-derived aortic phenotypes were obtained from approximately 39,000 UKBB participants from the European population (EUR)^4,9^. We sourced the SumStat for TAAD^10^ from the Million Veteran Program (MVP) for EUR (7,050 cases/ 330,610 controls), African (AFR) (1,266 cases/ 88,107 controls), and Hispanic (HIS) (310 cases/ 34,326 controls) populations. The GT exposure SumStats were derived from the population-specific GWAS of BMI adjusted- FG, FI, HbA1c, or 2hPG in EUR, AFR, and HIS^11^. The GT GWAS were performed in participants with no diabetes diagnosis, no reported use of diabetes-relevant medication(s); and who had an FG ≤ 7 mmol/L (126 mg/dL), 2hPG ≤ 11.1 mmol/L (200 mg/dL) or HbA1c ≤ 6.5%. IVs for T2DM were selected from EUR, HIS, and AFR-specific meta-analysis of T2DM adjusted on BMI^12,13^. To avoid sample overlap, we assess the association between T2DM and MRI-derived aortic phenotypes using EUR-only Sumstat which did not include the UKBB data. For the association between T2DM and TAAD, we employed T2D population specific SumStat for EUR, AFR, and HIS that did not include data from the Million Veteran Program. The GWAS sample size for GWAS for each trait in each population used in our MR is available in Table S1.

### Association analysis of aortic phenotypes and glycemic traits

First, we examined the epidemiological association of GT, including binary T2DM status, with aortic phenotypes including TAAD in the UKBB. We utilized blood glucose levels at enrollment, specifically random glucose (due to the small number of UKBB participants with both fasting glucose and MRI-derived aortic phenotypes) and HbA1c, and defined T2DM using International Classification of Diseases 10 (ICD-10) code, self-reported medical history and self-reported medication usage for diabetes (Table S2). The MRI-derived aortic measures included the diameter of the ascending aorta (AAoD), descending aorta (DAoD), as well as the distensibility of the ascending (AAoDis) and descending (DAoDis) aorta. These parameters were derived using image segmentation of the transversal image of cardiac MRI, sourced from an extensive dataset of over 1M images obtained from ∼39,000 UKBB participants^4,9,14^. Phenotyping of TAAD was defined using ICD-10 codes, OPCS codes, and self- reported medical history (Table S2).

The associations between GT and MRI-derived aortic phenotypes were assessed using multivariable linear regression models while the association between GT and TAAD was assessed using multivariable logistic regression. All analyses were adjusted on body surface area (BSA) at imaging, sex acquired from the central registry at recruitment, lipids level and blood pressure (measured at enrollment), and age at the time of imaging or at enrollment. To mitigate potential confounding by indication, adjustments were made for participants taking antihypertensive medication by adding 10 mmHg to diastolic blood pressure and 15 mmHg to systolic blood pressure^15^. For participants on lipid-lowering medications, raw LDL values were divided by 0.7 to account for medication influence on lipid levels^16^. Participants with diabetes medication usage were classified as T2DM cases.

### Mendelian randomization (MR) analyses Selection of instrumental variables (IV)

The IV was defined as SNP with minor allelic frequency (MAF) >0.05, independently associated with the exposure at the GWAS significance (p<5x10^-08^ and R^2^<0.001). However, to overcome power issues due to the small sample size in GWAS among AFR and HIS, we lowered the p-value threshold for selecting the IV to a suggestive threshold of p<1x10^-05^. In a second approach, we performed an MR using SNPs that were selected as IVs in EUR and that were nominally significant (p<0.05) in non-EUR. Variants selected as instruments from the exposure GWAS SumStat were then matched to the outcome SumStat. Given the lack of data available for aortic measures in non-EUR, we performed the MR of GT and aortic measurements among EUR-only, while the MR between GT and TAAD was performed independently in EUR, AFR, and HIS.

### Statistical analyses

Figure 1 presents the analysis flowchart. For each exposure-outcome pair, the IV was harmonized to ensure consistency of the effect allele between the exposure and the outcome. An IV was excluded if its variance, as explained by the F statistic, was less than 10 (F < 10 indicates a weak IV) if the MAF < 0.05, or if the SNP appeared to be multi-allelic. Additionally, we used MR-Steiger filtering to exclude SNPs indicating possible reverse causation or for which the effect on the outcome is larger than the exposure^17^. The inverse-variance weighted (IVW) method was used as the primary analytical approach to assess the relationship between each exposure-outcome pair. The IVW method is considered most powerful because it depends on the validity of all IVs and can robustly detect associations^18^. To investigate heterogeneity, we used the Cochran Q method. The MR-Presso global test was conducted to examine the presence of heterogeneous SNPs, identify outliers, and correct for horizontal pleiotropy^19^. In sensitivity analyses, we employed MR-Egger, weighted median, and inverse weighted median methods. The MR-Egger intercept test was specifically utilized to detect potential directional pleiotropy, where the MR-Egger intercept can be interpreted as an estimate of the average horizontal pleiotropic effect of the genetic variants ^20^.

**Figure 1:**
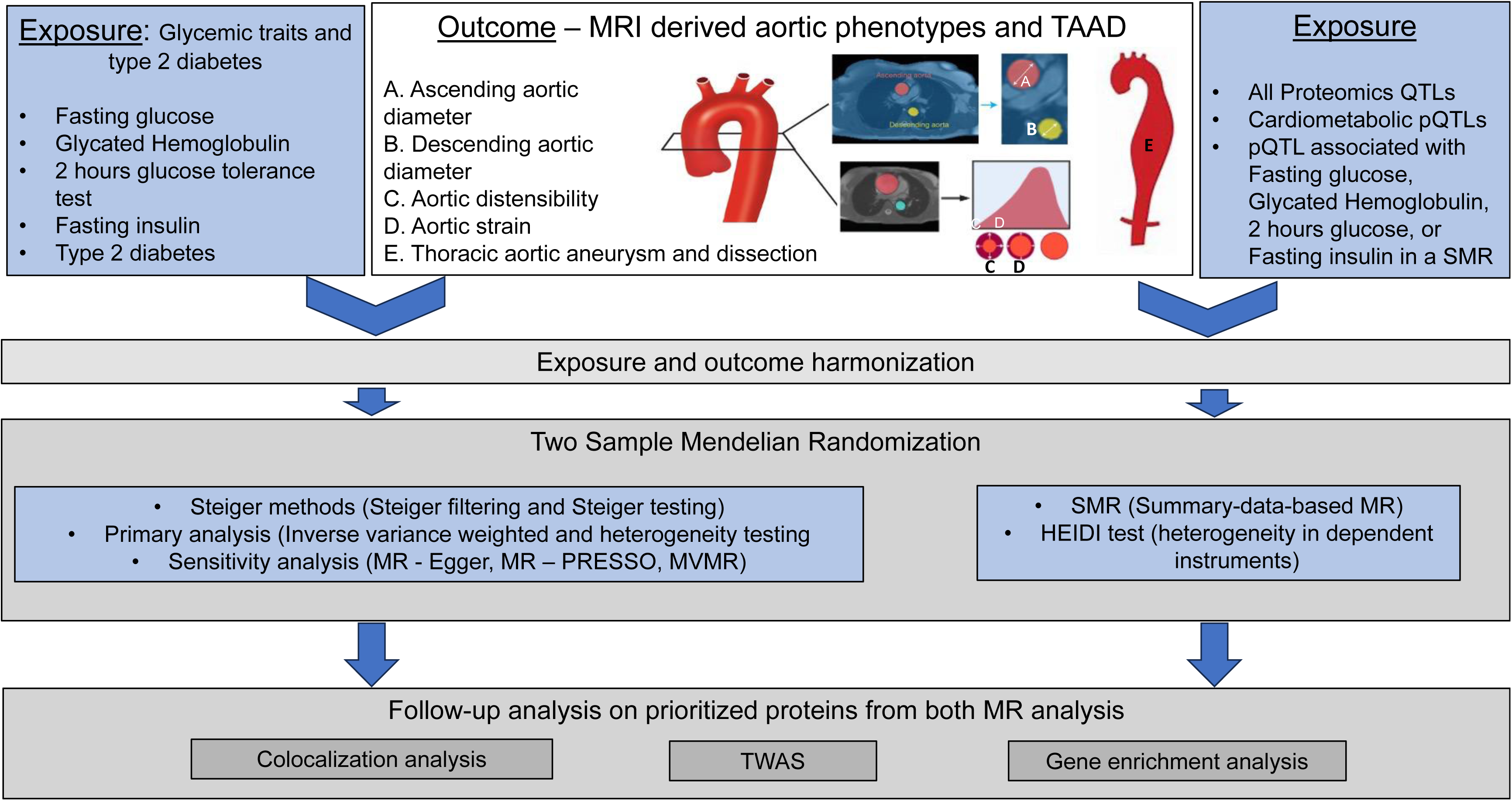
Flowchart Mendelian Randomization analysis investigating the causal association between glycemic traits and aortic measurement and diseases. Sample size description for each summary statistic is presented in the table S2.

To address the high likelihood of pleiotropy between GT genetic variants (that is, variants affecting multiple GT simultaneously), we conducted a multivariable MR analysis using MVMR in R ^21^. This analysis included all evaluated GTs (FG, FI, 2hPG, and HbA1c) and T2DM for each outcome. Additionally, we performed MVMR incorporating all evaluated GTs and T2DM, along with blood pressure (SBP, DBP) and lipid biomarkers (LDL, HDL) to account for potential confounders in the relation between GT and aortic phenotypes. For exposures and outcomes showing significant heterogeneity after pleiotropy assessment, reverse causation testing and outlier correction, we employed a clustering approach with MR-Clust ^22^. MR-Clust clusters genetic variants based on their causal effect estimates and define clusters assuming distinct causal mechanisms.

To explore the potential biological mechanisms relevant to T2DM driving the observed inverse association with aortic phenotypes, we leveraged the T2DM mechanistic clusters defined by Suzuki et al^13^ for MR. These T2DM mechanistic clusters were derived using a combination of dimension reduction and hard clustering of T2DM lead SNPs and their significant association with cardiometabolic traits such as FG, HbA1c, BMI, obesity, blood pressure, lipids biomarkers, and fat tissue percentage^13^. The defined clusters include Beta cell +PI (Proinsulin; 91 SNPs), Beta cell-PI (89 SNPs), Residual glycemic (389 SNPs), Body fat (273 SNPs), metabolic syndrome (166 SNPs), obesity (233 SNPs), lipodystrophy (45 SNPs), liver and lipid metabolism (3 SNPs). For each of these clusters, we performed an MR analysis with MRI-derived aortic phenotypes and TAAD.

### Evaluation of the MR assumption

Overall, to ensure that robust conclusions are derived, we rigorously tested the three core MR assumptions: relevance, exchangeability, and exclusion restriction. The relevance assumption was validated by assessing the strength of IV by excluding instruments with an F statistic below 10 to avoid weak instrument bias. The exchangeability assumption was addressed through multivariable MR analysis^21^ to distinguish genuine gene-disease associations from spurious ones caused by confounding genetic variants^23^. Finally, the exclusion restriction assumption was examined using MR-Steiger filtering^17^, which helps confirm that the genetic predictors influenced the outcome solely through exposure.

### Mendelian Randomization of pQTLs and GT as well as aortic phenotypes

To investigate the colocalization of protein quantitative trait loci (pQTLs) and GT as well as aortic phenotypes, we employed summary-data-based Mendelian Randomization (SMR) and the heterogeneity in dependent instruments (HEIDI) test^24^. GWAS of 2,940 plasma proteins were sourced from the genetically inferred EUR in the UK Biobank^25^. *cis*-pQTLs were defined as SNPs located 1 Mb upstream and downstream of the protein-coding genes, demonstrating an association with plasma protein levels at P < 3x10^-05^. Multi-allelic SNPs were excluded before the SMR analysis. We conducted SMR analysis integrating GWAS with pQTL data from the UKBB with GWAS of GT, T2DM and aortic phenotypes from EUR. The EUR population in the 1000 Genomes Project was used as the reference to estimate the linkage disequilibrium between SNPs. The HEIDI test was then applied to differentiate pleiotropic associations from those due to linkage. Associations passing the Bonferroni-corrected significance threshold in SMR analysis and the HEIDI P > 0.05 were considered robust evidence for colocalization between pQTLs and GWAS.

### Functional annotation of IVs and transcriptome-wide association studies

To explore the biological implications of GT genetic variants, we first annotated each SNP selected as IV using various annotation tools including Phenoscanner^26^, SNPNexus^27^, and Haploreg^28^. These annotations include eQTL expression look-up for each SNP in various tissues (such as aorta, arteries, heart, and fibroblasts), with eQTL data sourced from GTEx and STARNET^29,30^. A significant eQTL mapping was defined as an SNP with tissue eQTL p-value < 1x10^-05^. To further extend our eQTL mapping to genes targeted by our IVs, we performed a transcription-wide association (TWAS) study on all GT traits GWAS using S-PrediXcan, a summary-statistics-based gene mapping method available in the MetaXcan software package^31^. The summary statistics were harmonized following the best practices on MetaXcan GitHub (https://github.com/hakyimlab/MetaXcan). Using a precomputed expression prediction model utilizing the GTEx database, we inferred tissue-specific gene-trait associations^30^. Tissues tested include coronary artery, aorta, visceral and subcutaneous adipose, atrial appendage, left ventricle, liver, pancreas, whole blood, and EBV-transformed lymphocyte cells. In sensitivity analyses, we used two different families of prediction models, elastic net-based and MASHR-based, a biologically informed model^32^. For each trait, FDR p-value correction was conducted both by tissue and across all tested tissues.

### Gene enrichment analysis

Significant genes from S-PrediXcan that were also gene targets by the IVs in our MR, were tested for GO-term overrepresentation using the enrichGO function within the clusterProfiler package^33^. The analysis was performed for each ontology (molecular function, biological process, and cellular component) and significant genes from each trait-tissue pair separately. The universe of genes for each enrichment analysis was defined as genes present in the GTEx tissues summary statistic files. The p-values were corrected using the Benjamini-Hochberg method.

## RESULTS

### Epidemiological association between GT and aortic phenotypes in the UKBB

The characteristics of our study population are described in Table ST2. We observed an inverse association of HbA1c and T2DM with ascending and descending aortic diameters as well as TAAD risk (Table 1). The inverse association with HbA1c was even stronger among participants without diabetes (Table 1, Table ST4).

### MR Glycemic traits and aortic phenotypes Aorta diameter and thoracic aortic aneurysm

In univariate MR analyses using the IVW method, 2hPG, FG, HbA1c, and T2DM were all inversely associated with AAoD and DAoD but no association was observed with FI (Table S5-6, Figure 2). Notably, the associations of 2hPG, FG, HbA1c, and T2DM with AAoD were significant (Table S6, Figure 2), whereas only the associations of 2hPG and FG with DAoD were significant (Figure 2). We also observed a significantly inverse association of 2hPG, FG, HbA1c, and T2DM with TAAD (Table S6, Figure 2). These observed association with TAAD in Europeans were similarly present among AFR and HIS populations (Figure S3, Table ST6). However, only the association with T2DM in HIS reached statistical significance (β: -0.35, p=7x10^-03^) (Table ST6, Figure S3). In sensitivity analyses, a consistent inverse association was observed using MR Egger, weighted median, and weighted modal approaches, which aligned with the aforementioned IVW results (table S5-6, Figure S2).

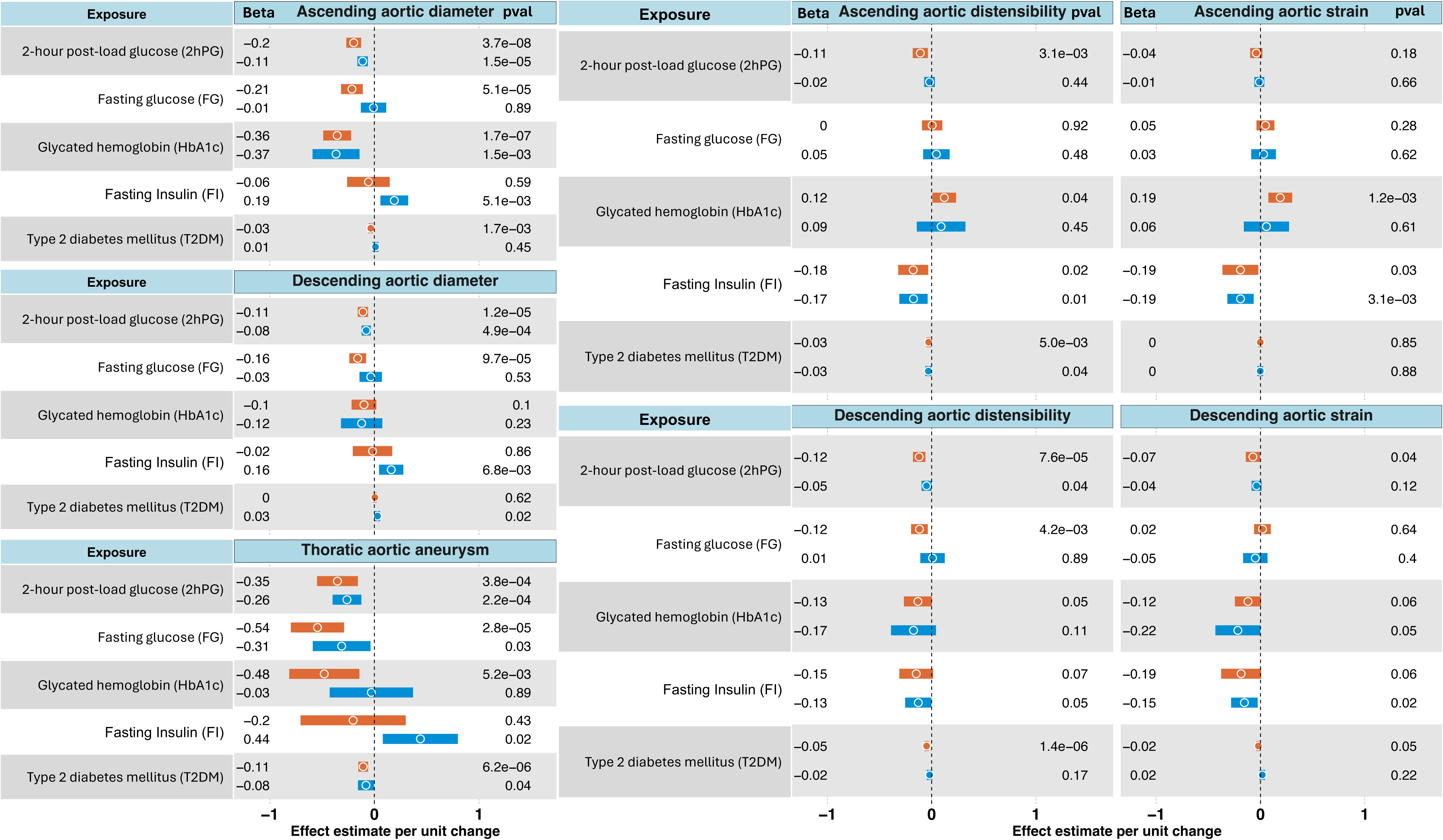

In multivariable MR analyses, the inverse associations remained significant for 2hPG and AAoD (β: -0.11, p=2x10^-5^); HbA1c and AAoD (β: -0.37, p=4x10^-03^); 2hPG and DAoD (β: -0.08, p=5x10^-04^); 2hPG and TAA (β: -0.27 (OR=0.76), p=2.3x10^-05^); FG and TAAD (β: -0.26 (OR=, p=3.5x10^-02^); and T2DM and TAAD (β: -0.07, p=2x10^-02^) (Figure1, Table ST7). It is noteworthy that some exposures had weak strength of the genetic instruments (F-statistics <10, Table ST7) due to a large number of SNPs used as IVs in the T2DM analysis. In a sensitivity analysis using a different summary statistic with smaller number of genetic instruments for T2DM, we observed similar results with most exposures displaying strong genetic instruments (Table ST8).

### Aortic strain and distensibility

In univariate MR analyses using IVW, we observed that 2hPG exhibited an inverse association with both DAoDis and AAoDis (β = -0.12, p = 7.5 x 10^-5^ and β = -0.11, p = 3.1 x 10^-3^, respectively; see Figure 2). FG and T2DM were significantly associated with DAoDis (β = -0.12, p = 4 x 10^-3^ and β = -0.03, p = 5 x 10^-3^ for FG and T2DM, respectively). Regarding aortic strain (AoSt), FI and HbA1c demonstrated a significant association with strain in the ascending aorta (Figure 2). In multivariable MR analyses, the association between 2hPG and DAoDis remained significant, and the associations between AAoSt and either FI or HbA1c also persisted (Figure 2).

### Heterogeneity and Sensitivity Analysis

Significant variability was detected using the Q statistic for the associations between DAoD and HbA1c; TAAD and T2DM; and FI with both AAoD and DAoD (Table ST5), despite addressing reverse causation, excluding weak instruments, correcting for horizontal pleiotropy and removing outliers. However, consistent directionality of effects was observed across MR IWV, MR Egger, weighted median, and weighted modal approaches for most exposure-outcome traits (Table ST5, Figure S2-4).

### MR using the clustering approach

Given that the observed heterogeneity suggests a potential contradiction in the effect direction of the SNPs included in each analysis, we reclassified our IVs into more homogenous groups using a cluster analysis approach and conducted MR within each cluster. We identified, for the outcome AAoD, DAoD, and TAAD, and the exposure HbA1c, FG, and FI respectively, two distinct clusters of associations (Table ST9, Figure 3(a)). These clusters included either one with a positive association and another with a negative association, or two unique clusters with negative associations. For instance, the inverse relationship between FG and AAoD showcased two clusters with inverse associations, of which only one cluster, comprising 20 IVs, was significant (β=-0.88, p=8.02x10^-20^, Figure 3(a)) while the inverse association with the second cluster including 48 IV was not significant (Table ST9. Figure 3(a)). The associations between FI and both AAoD and DAoD, which were non-significant in our overall MR analysis, revealed two distinct clusters with significant associations. One cluster displayed a positive association, while the other showed an inverse association (Figure 3a). This indicates that a subset of insulin-related genetic instruments may contribute to an increase in aortic diameter, while another set may be linked to a decrease in aortic diameter. Similarly, we observed two distinct clusters with significant associations in opposite directions for FG/TAAD and T2DM/TAAD exposure-outcome pairs. This suggests that the impact of FG or T2DM on TAAD may operate through distinct biological pathways, which are either protective or detrimental.

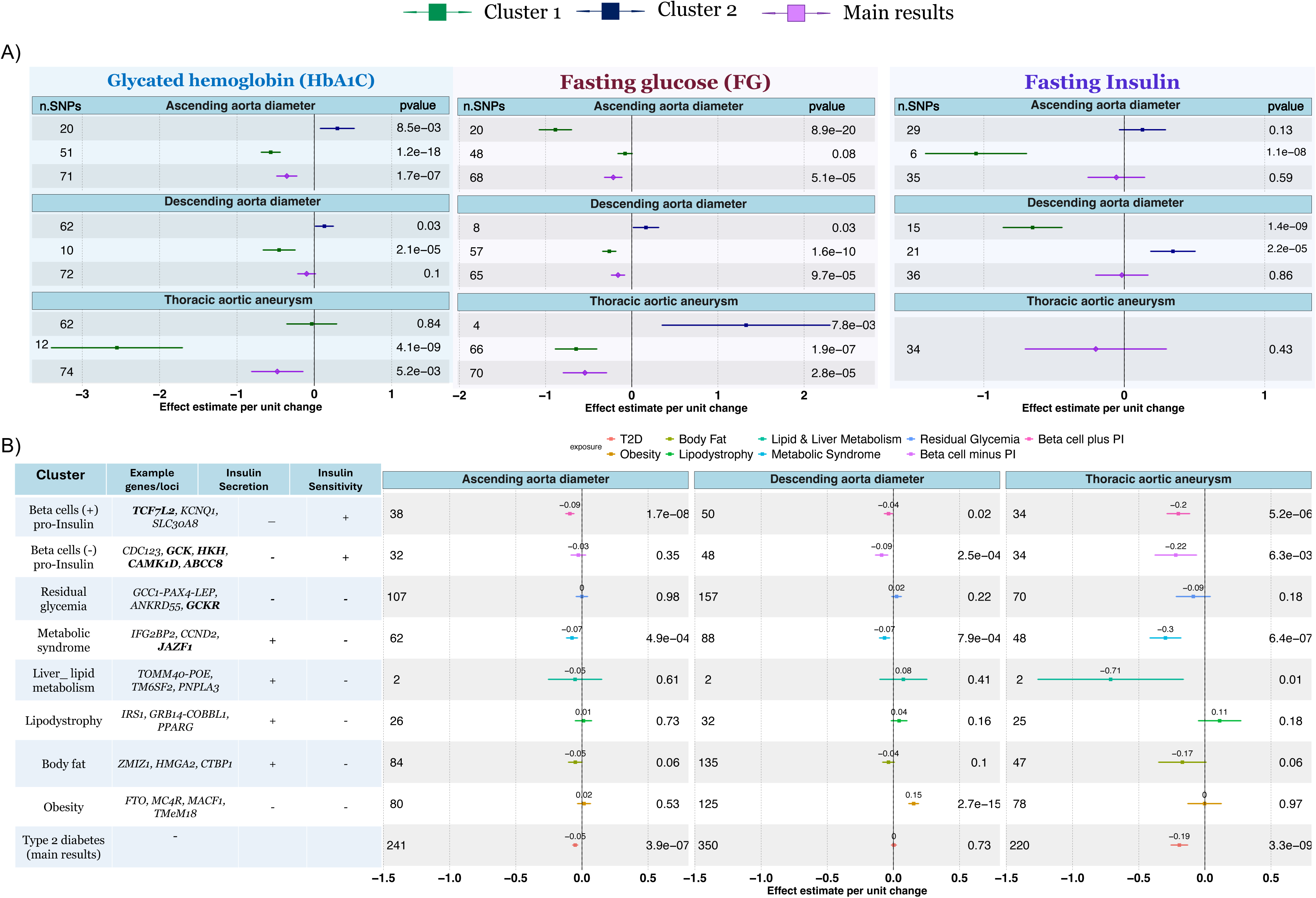

### MR using T2DM predefined cluster

To further explore the biological processes linking T2DM to MRI-derived aortic phenotypes and TAAD, we conducted MR between aortic phenotypes and clusters of SNPs representing various biological processes implicated in T2DM pathophysiology, as recently described by Suzuki et al ^13^. We found that the beta-cell (+) proinsulin cluster (beta-cell +PI) and the beta-cell (-) proinsulin (beta-cell -PI), characterized by variants that reduce insulin secretion and enhance insulin sensitivity, were significantly inversely associated with AAoD, DAoD, and TAAD (Figure 3b, Table ST10). These associations were most pronounced for the beta-cell -PI clusters. Furthermore, the metabolic syndrome cluster (dominated by fasting glucose, fasting Insulin, visceral adipose tissues, and Glutamine fructose-6-phosphate amidotransferase [GFAT] genes) was inversely associated with AAoD, DAoD, and TAAD, while the obesity cluster was significantly associated with larger DAoD (Figure 3b, Table ST10). Additionally, the lipodystrophy cluster, enriched with body fat, lipid levels, and blood pressure, were associated with an increase in AAoD, DAoD, and TAAD (although the association was not significant), while decreasing strain and distensibility of both ascending and descending aorta (Figure 3b and Figure S5).

### Proteomic mendelian randomization

To better understand the link between glycemic traits and aortic phenotype, we also assessed the impact of proteomic biomarkers on aortic structure and diseases. We performed SMR between UKBB 2940 Olink pQTL and aortic phenotypes and identified few pQTL associated with aortic phenotypes (Table ST13, Figure 4a and Figure S6). For instance, COL6A3 pQTL was inversely associated with AAoD, DAoD, and TAAD (Figure 4a). *FGF5* pQTL was associated with AAoD and AAostr, AGER, ECM1, and IGFBP3 were associated with DAoD, DAoStr, and DAdis respectively (Figure 4a). Other pQTL with significant SMR association includes *FADD, GLRX,* and *AOC3* which are all pQTL biomarkers for cardiometabolic profile (Table ST13). To investigate whether pQTLs associated with GT are also associated with aortic phenotype, we performed SMR analysis for FG, 2hPG, FI, and HbA1c. We then checked for overlap between pQTLs showing significant SMR associations with both aortic phenotypes and GT. We identified 12 proteins associated with HbA1c, 4 proteins associated with FG, 3 proteins for FI, and one protein for 2hPG (Table ST13). Among these identified proteins, only ABO which was associated with 2hPG, was also associated with DAoSt (Table ST13). No other proteins with significant association with GT were also associated with aortic phenotypes. When investigating whether SNPs in these pQTL-associated genes have IVs in our MR, we found that SNPs in or near *ABO* and *AGER* were present in the UVMR.

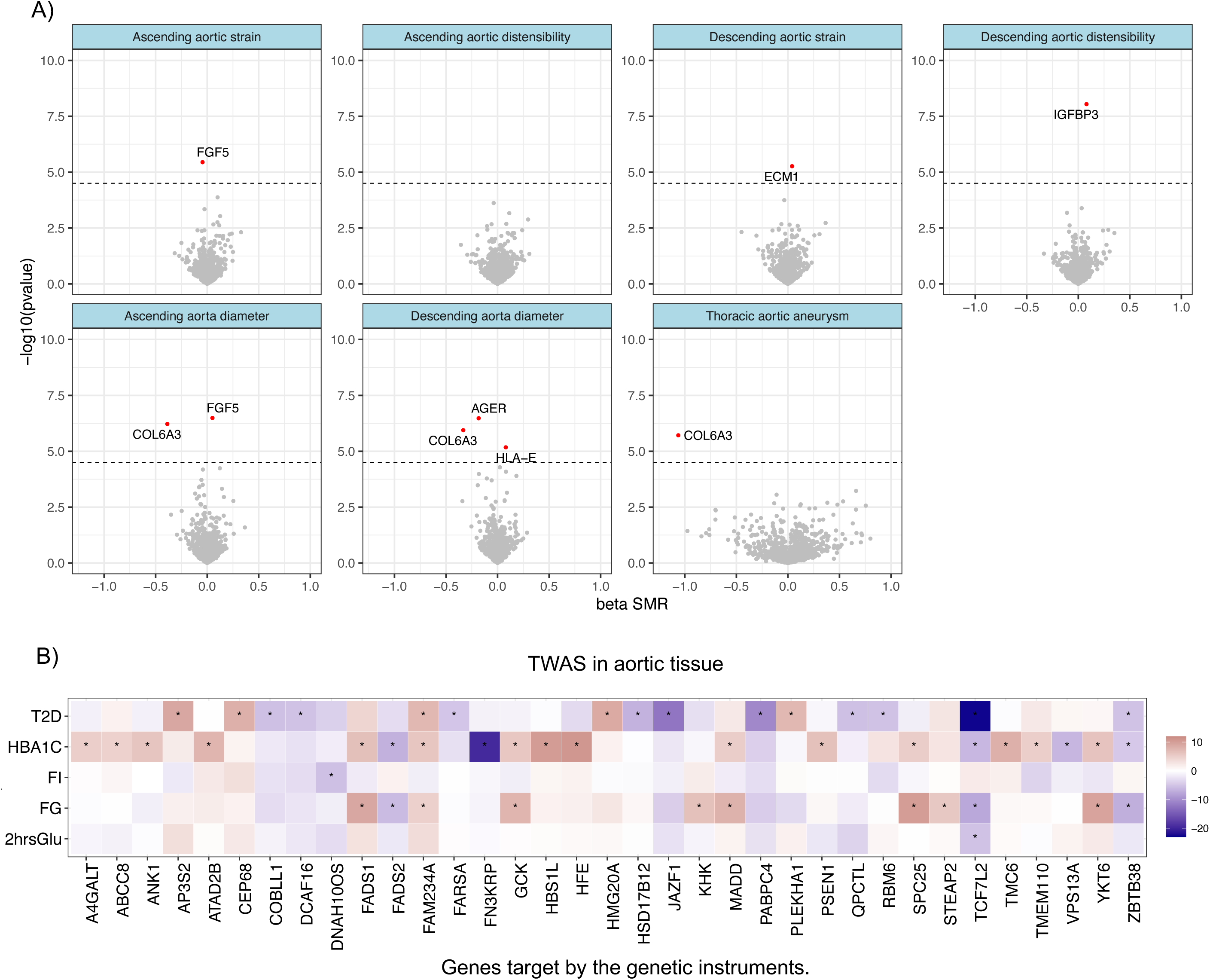

### Functional annotation and enrichment Analysis of Selected Genetic Instruments

Functional annotations of SNPs selected as IVs are available in Table S10 and S16. Overall, we observed that several of our IVs are located in regulatory regions such as promoter/enhancer histone marks, proteins bound, or altered transcription factors binding motifs. Several of our IVs were also missense (ex: rs267738 in *CESR2*, rs120326 in *GCKR*, and rs1800562 in *HFE*), nonsynonymous variants or variants located in 5’UTR of genes. We also observed that several IVs have significant eQTL changes in various heart-related tissues including the aorta (Table S13). For instance, SNPs selected as IVs in *GCK*, *TCF7L2*, *HFE, ABCC8, and AGER,* showed significant eQTL in heart and aortic tissues (Table S13). Our TWAS analysis unveiled a significant enrichment (p<1x10^-05^) of genes surrounding our IVs for FG, HbA1c, and T2DM across a range of tissues and cells including the aorta, tibial artery, heart, whole blood, and fibroblasts (Table ST13). When specifically examining the TWAS in aortic tissues, we observed a significant association between various GT and changes in gene expression of *GCK, CTSS, HFE, KHK, AGER, and TCF7L2* (Figure 4(b) and Table ST14). Moreover, our gene ontology enrichment utilizing gene targets by our IVs and with notable eQTL expression in aortic tissues showcased a significant enrichment in gene ontology pathways such as carbohydrate kinase activity, glucose homeostasis, intracellular glucose homeostasis, and positive regulation of hormone secretion (Table ST15). Genes playing a key role in these pathways include *TCF7L2, GCK, HFE, KHK, and AGER*.

## DISCUSSION

In this study, we comprehensively assessed the relationship between glycemic traits and aortic phenotypes, using an epidemiological approach, followed by a comprehensive MR across multiple populations, and a multi-omics analysis. Genetic predictors associated with high fasting glucose, HbA1c, 2-hour post-load glucose, and T2DM risk were linked to a decrease in aortic size and a reduced risk of TAAD. In particular, the inverse association with T2DM may be driven by genetic variants associated with beta-cell function. Moreover, glucose regulation genes such as *GCK* and *TCF7L2* are potentially involved in biological processes conferring a protective effect on aortic aneurysm.

Our study confirmed and expanded the inverse epidemiological associations between glycemic measures and aortic phenotypes observed in prior research. For instance, increased arterial stiffness, reduced aortic elasticity, and lower rates of development, progression, and mortality from aortic aneurysms have been noted individuals with prediabetes or diabetes^34–36^. Furthermore, among men, an inverse relationship has been observed between high fasting glucose levels and infra-aortic diameter on one hand, and the risk of abdominal aortic aneurysm (AAA) progression one the other hand^37^. Our MR analyses indicated that genetically predicted T2DM risk, higher HbA1c, FG, and 2hPG levels are associated with smaller AAoD, DAoD, and lower risk of TAAD; consistent with previously reported MR studies that examined the impact of GT on TAAD and MRI-derived aortic structure^38,39^. We also showed, for the first time, an inverse association between GT and TAAD among African and Hispanic populations. However, the association was only significant for T2DM in a Hispanic subgroup, probably due to a lack of power driven by insufficient sample size in ancestry-specific summary statistics.

An innovative aspect of our analysis is the application of MR on clusters of genetic variants, which allowed testing the hypothesis of the co-existence of different directional effects within our GT instruments, attributable to distinct biological and molecular pathways. Our approach revealed that certain IVs associated with FG, FI, and HbA1c may correlate with an increase in aortic diameter and TAAD risk. Conversely, other IVs associated with these GTs, were associated with a decreased in aortic diameter and TAAD risk. For instance, within the cluster of genetic variants showing an inverse association for FG and HbA1c, we identified variants in genes such as *GCK* and *TCF7L2*. Indeed, prior studies have suggested that variants in *TCF7L2* and *GCK* are linked to a decreased risk of aortic aneurysm or macrovascular disease^10,40,41^. On the other hand, among variants in the FI cluster associated with a larger aorta diameter or higher risk of TAAD, we noted the variant rs284585 in *VEGF*, a vascular endothelial growth factor. *VEGF* is a well-known protein that causes vascular endothelial cells to proliferate, migrate, and become more permeable^42^. Moreover, findings suggest that insulin’s action may enhance angiogenesis through *VEGF* and promote vasodilation^43^ thus possibly increasing the risk of TAAD.

We also utilized the cardiometabolic clusters defined by Suzuki et al ^13^ to investigate the relationship between various aspects of the T2DM metabolic profile and aortic phenotypes. We showed that clusters associated with beta-cell dysfunction—specifically those with a positive association with proinsulin (beta cell (+) PI) and a negative association with proinsulin (beta cell (-) PI)—were inversely correlated with aorta diameter and TAAD. Notably, the beta cell (-) PI cluster exhibited even stronger associations. Of note, the beta cell cluster was dominated by genes/loci increasing FG, 2hPG, and HbA1c suggesting that factors affecting glucose homeostasis have a greater impact on aortic structure and function.

We enhanced our MR analysis by additionally conducting a proteomic MR approach, which offers additional insights into proteins involved, and thus potential mechanisms linking GT to aortic structure and disease. This approach helped shed light on the biological pathways contributing to observed genetic associations, thereby strengthening our findings and providing a comprehensive perspective on the molecular underpinnings of aortic phenotypes. Our SMR analysis identified several proteomic biomarkers associated with aortic phenotypes. Notably, *COL6A3* pQTL is associated with AAoD, DAoD, and TAAD while AGER pQTL is associated with DAoD. AGER or RAGE is a receptor that binds to advanced glycation end products (AGEs) for degradation. Chronic hyperglycemia leads to the formation of AGEs, which stabilize collagen networks, increase resistance to protease degradation, and reduce aortic wall stress^7,43–45^. In AAA tissues from individuals with diabetes, increased cross-linking AGEs like pentosidine correlate with smaller AAA diameters, indicating a protective role due to collagen network stabilization^7,46^. COL6A3 is a core component of collagen type VI, which assembles into microfibrils, forming a scaffold within the extracellular matrix, thus supporting the aortic tissue structure by providing tensile strength and resilience against mechanical stress. COL6A3 can undergo glycation in the setting of hyperglycemia. AGE-modified COL6A3 in the aortic wall could contribute to increased vascular stiffness, influencing blood flow dynamics and vessel function^47–49^. Further studies are needed to better understand the relation between these protein biomarkers and aortic phenotype in hyperglycemic conditions.

Functional annotation of IVs revealed an enrichment of functional impact for multiple SNPs. this includes the presence of missense variants among our IVs, SNPs located in regulatory elements such as transcription factor binding sites, enhancers, and promotors active in aortic tissues. This includes SNPs in genes such as *GCK* that also show significant enrichment in aortic tissues from our TWAS. *GCK*, a glucokinase linked to maturity-onset diabetes of the young (MODY2), a rare monogenic diabetes where elevated blood glucose levels remain stable without worsening glycemic control^41,50^. Studies have suggested that patients with GCK- MODY2 exhibit a lower risk of diabetes-related micro- and macro-vascular complications^41^. Glucokinase activation, targeting *GCK*, has recently emerged as a potential diabetes therapy, which is either FDA approved (tofogliflozin) or tested in phase II or III clinical trials (ex: GKA-50 and dorzagliatin)^51–54^. In our study, SNPs in *GCK* were included as IVs for the HbA1c, FG, and the T2DM beta cell (-) PI cluster. Molecules targeting *GCK* in diabetes treatment could be good candidates for repurposing to TAAD treatment and prevention given the impact of *GCK* on decreasing the aorta diameter.

Another interesting finding provided by MR, functional annotation and TWAS analysis in aortic tissue is the evidence to support the causal effect of *TCF7L2* gene on adverse aortic phenotypes. An inverse association between *TCF7L2* locus and TAA was previously described by Roychowshury et al^40^. However, in the latter study, the inverse association observed at the locus was driven by a set of variants also independently associated with an increased risk of diabetes. They suggested that the inverse association between T2DM and TAAD at this locus reflects independent gene-level horizontal pleiotropy^40^. In our TWAS, *TCF7L2* gene expression in aortic tissues was inversely associated with HbA1c, FG, 2hPG, and T2DM. Furthermore, SNPs in *TCF7L2* including rs7903146 were IVs for 2hGP, HbA1c (-) association cluster, FG (-) association cluster as well as in the T2D beta cell (+) PI cluster. Indeed, rs7903146 is a known diabetes candidate SNP^55^. In our analysis, rs7903146 showed an inverse association with 2hPG, FG, HbA1c, and T2DM with the directionality of the effect consistent with the association of TCF7L2 variants rs4077257 reported by Roychowshury et al^40^. A genetic correlation analysis conducted on a European population revealed a correlation coefficient (R²) of 0.46 between the genetic variants rs7903146 and rs4077257. Based on these findings, we hypothesize that the observed inverse association between the *TCF7L2* gene and aortic phenotypes may be mediated through glucose levels, independent of diabetes status. Further research is warranted to investigate this hypothesis in greater detail.

Our pathway analyses identified several pathways with high enrichment of genes targeted by the IVs in our MR study. These pathways include carbohydrate kinase activity, glucose homeostasis, response to intracellular glucose homeostasis, response to monosaccharides, regulation of the glycolytic process, peptide secretion, and response to carbohydrates. Key genes involved are *GCK*, *TCF7L2*, *KHK*, *HFE*, and *AGER*. These interconnected pathways are crucial for maintaining glucose balance and cellular energy management. For example, carbohydrate kinase activity and glucose homeostasis regulate glucose levels and limit AGE formation. As mentioned above, accumulation of AGEs affects the extracellular matrix remodeling of the aortic wall by cross-linking with ECM proteins such as collagen and elastin which could increase aortic wall stiffness^7^. Thus, possibly contributing to the observed decreased risk of aortic aneurysm in diabetes and hyperglycemia.

### Strengths and limitations

To our knowledge, our study is the first to investigate the role of GT on TAAD among non-European populations. Furthermore, this is the first comprehensive study to assess the relationship between GT/diabetes and aortic phenotypes using a multi-omic approach that includes MR, clustering MR analysis, proteomic MR, TWAS, and gene/pathway enrichment analysis. The validity and robustness of our findings are supported using a two-sample MR technique with the latest GWAS summary statistics data available for exposure-outcome traits, supplemented by MR cluster analysis. A limitation of our study is that we could not assess the causal relationship between glycemic traits and aortic structure in non-European populations. However, our results showing an inverse association between glycemic traits and thoracic aortic aneurysm suggest that the direction of these associations is consistent across different ancestries.

## CONCLUSION

Genetic predictors associated with high fasting glucose, HbA1c, 2-hour post-load glucose, and T2DM risk are causally linked to a decrease in aortic size and a reduced risk of TAAD. The inverse association with T2DM may be driven by genetic variants associated with beta-cell function. The study highlights the potential involvement of glucose regulation genes such as *GCK* and *TCF7L2*. Hence, drug targeting GCK such as glucokinase activators, or interacting with TCF7L2 such as metformin, could be potential good candidate to be repurposed for treating and preventing TAAD. Future preclinical/clinical research is needed to better understand this opportunity.

## Data Availability

All data produced in the present study are available upon reasonable request to the authors

## Notes

### Competing Interest Statement

We thank the Type 2 Diabetes Global Genomics Initiative (T2DGGI) for generation of association summary statistics without UKBB and MVP; H.T. is supported by the Gates-Cambridge Trust and NIH Oxford-Cambridge scholars program;MGL was supported by the Doris Duke Foundation (Award 2023-0224) and US Department of Veterans Affairs Biomedical Research and Development Award IK2-BX006551, and has received research funding to the institution trial from Myome unrelated to this work. The content of this manuscript does not represent the views of the Department of Veterans Affairs or the United States Government; D.K. Is an employee at and has equity in Bitterroot Bio unrelated to the current research;

### Funding Statement

CT received K01 award and AHA funding support

### Author Declarations

The study used openly available before initiation of the study and original data sources are stated in the supplementary material of the manuscript with the accession number of publications with original data sources

