## Supplementary_material for "Mendelian randomization suggests a causal link between glycemic traits and thoracic aortic structures and diseases"

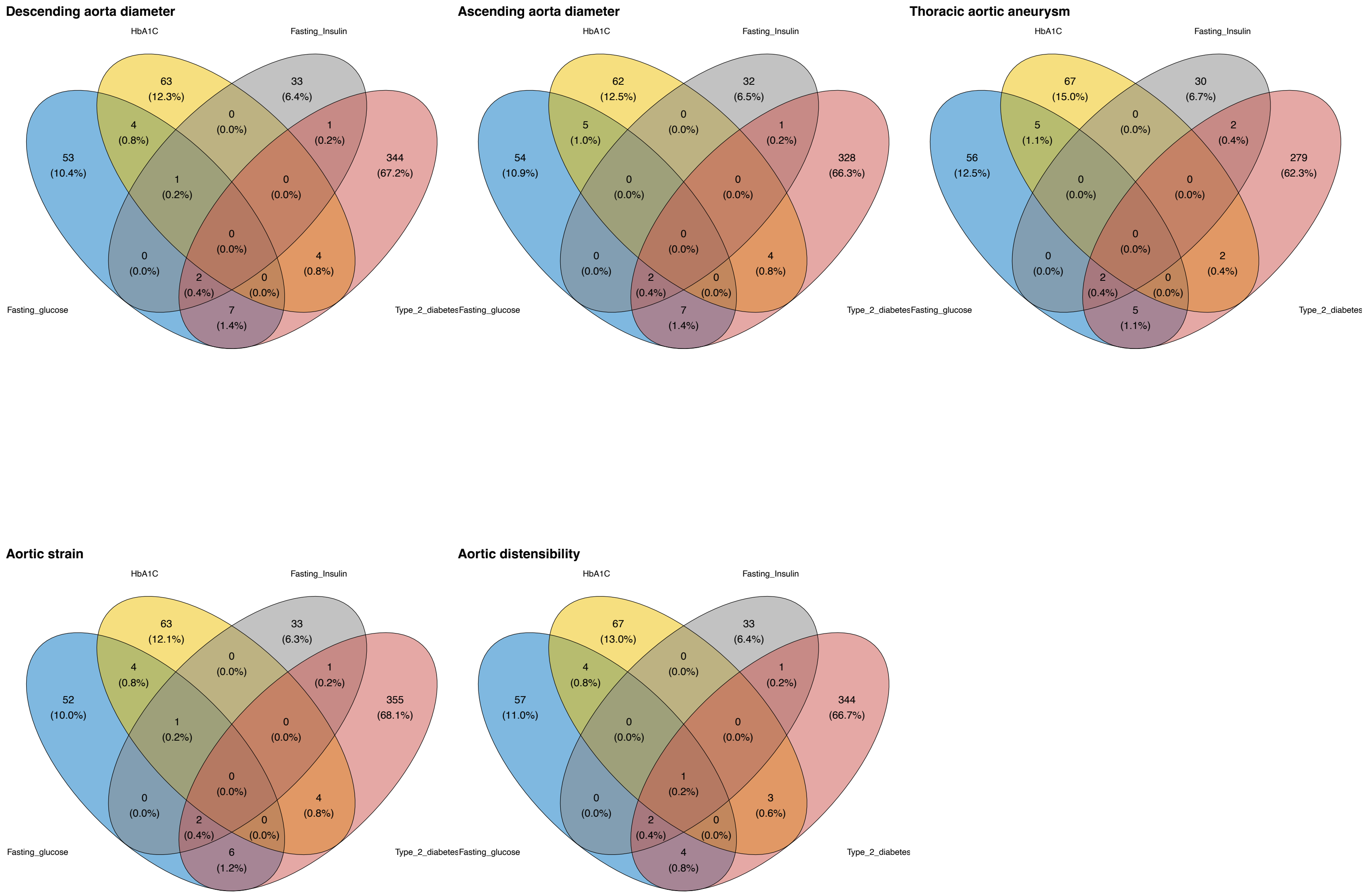


Figure S1: Venn Diagram genetic instruments

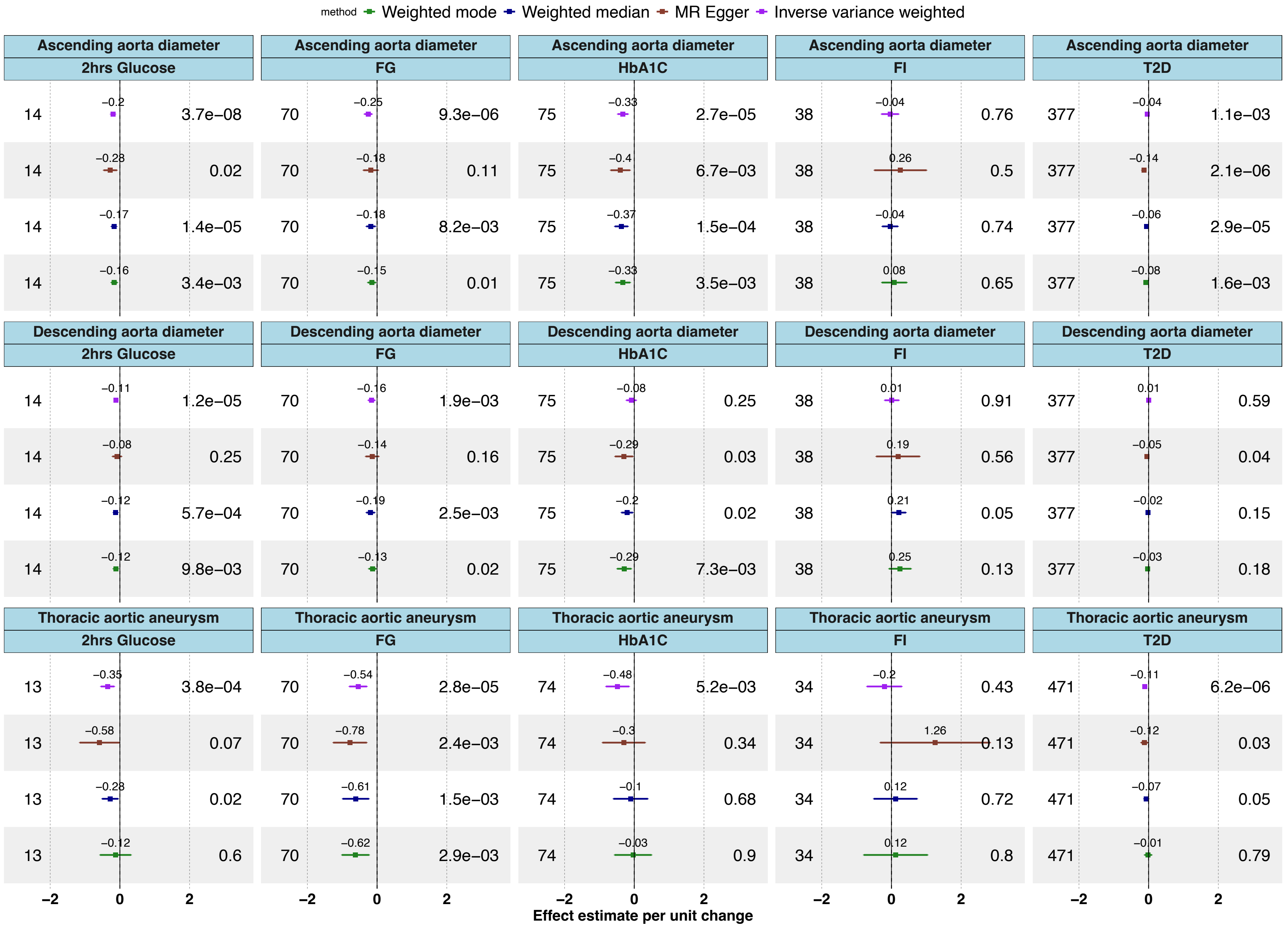


Figure S2: Univariate MR, heterogeneity test and sensitivity analysis for AAoD, DAoD, and TAAD

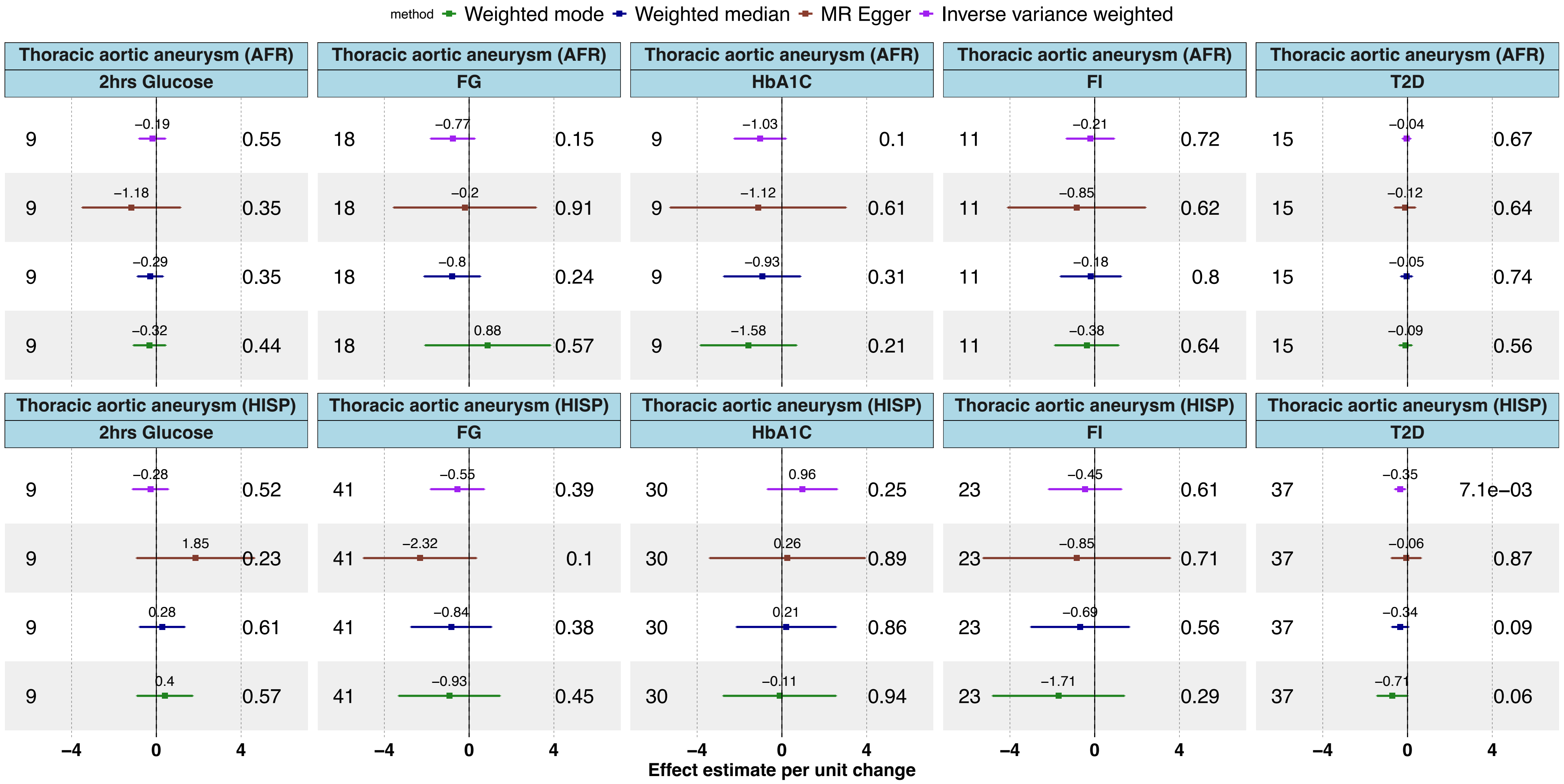


Figure S3: Univariate MR, heterogeneity test and sensitivity analysis for TAAD in Hispanics (HISP) and African (AFR) ancestries

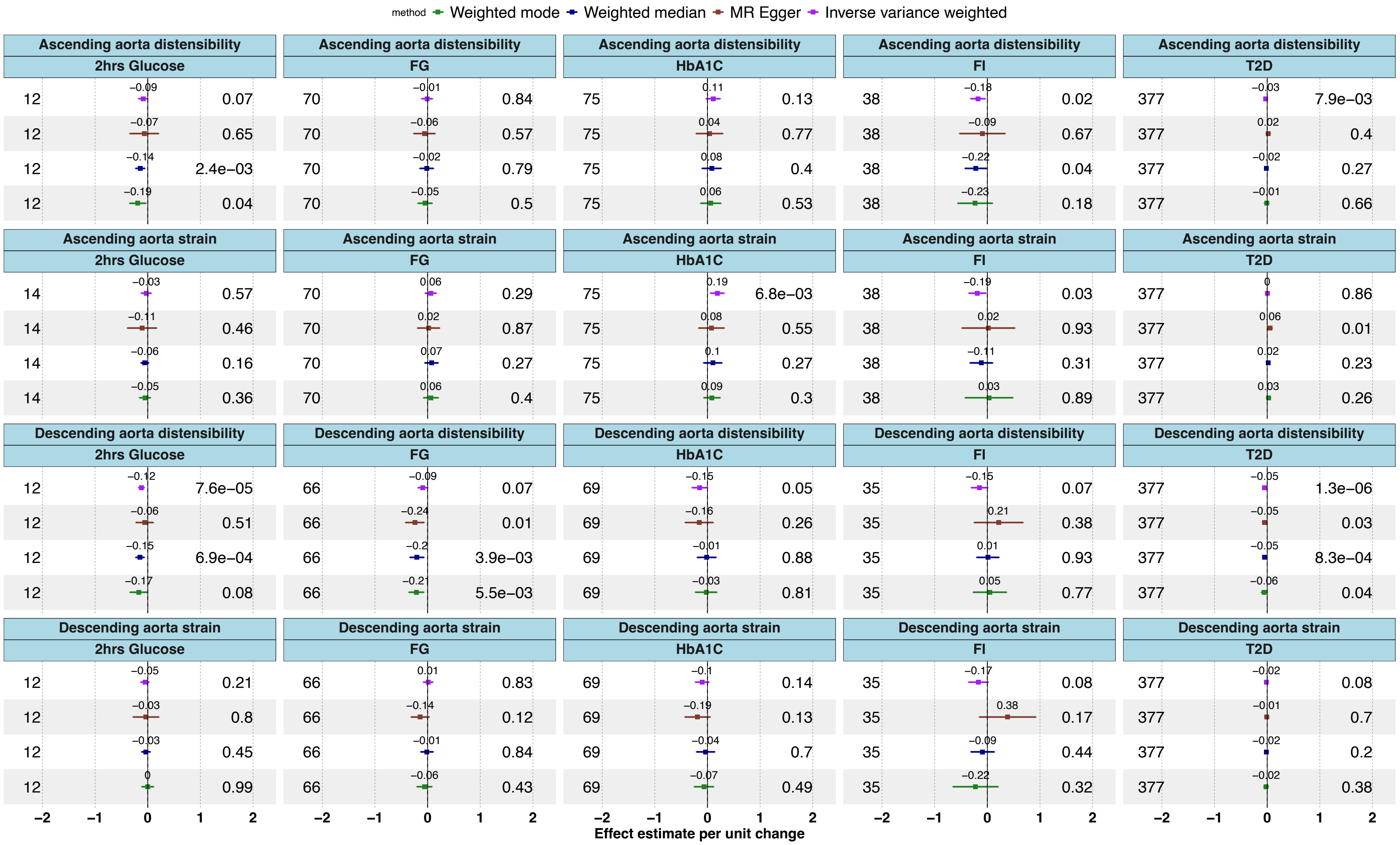


Figure S4: Univariate MR, heterogeneity test and sensitivity analysis for ascending and descending aorta strain and distensibility

| Cluster | Example genes/loci | Insulin Secretion | Insulin Sensitivity |
|---|---|---|---|
| Beta cells (+) pro-Insulin | ***TCF7L2***, *KCNQ1, SLC30A8* | – | + |
| Beta cells (-) pro-Insulin | *CDC123*, ***GCK***, ***HKH***, ***CAMK1D***, ***ABCC8*** | - | + |
| Residual glycemia | *GCC1-PAX4-LEP, ANKRD55*, ***GCKR*** | - | - |
| Metabolic syndrome | *IFG2BP2, CCND2*, ***JAZF1*** | + | - |
| Liver_ lipid metabolism | *TOMM40-POE, TM6SF2, PNPLA3* | + | - |
| Lipodystrophy | *IRS1, GRB14-COBBL1, PPARG* | + | - |
| Body fat | *ZMIZ1, HMGA2, CTBP1* | + | - |
| Obesity | *FTO, MC4R, MACF1, TMeM18* | - | - |
| Type 2 diabetes (main results) | - | | |

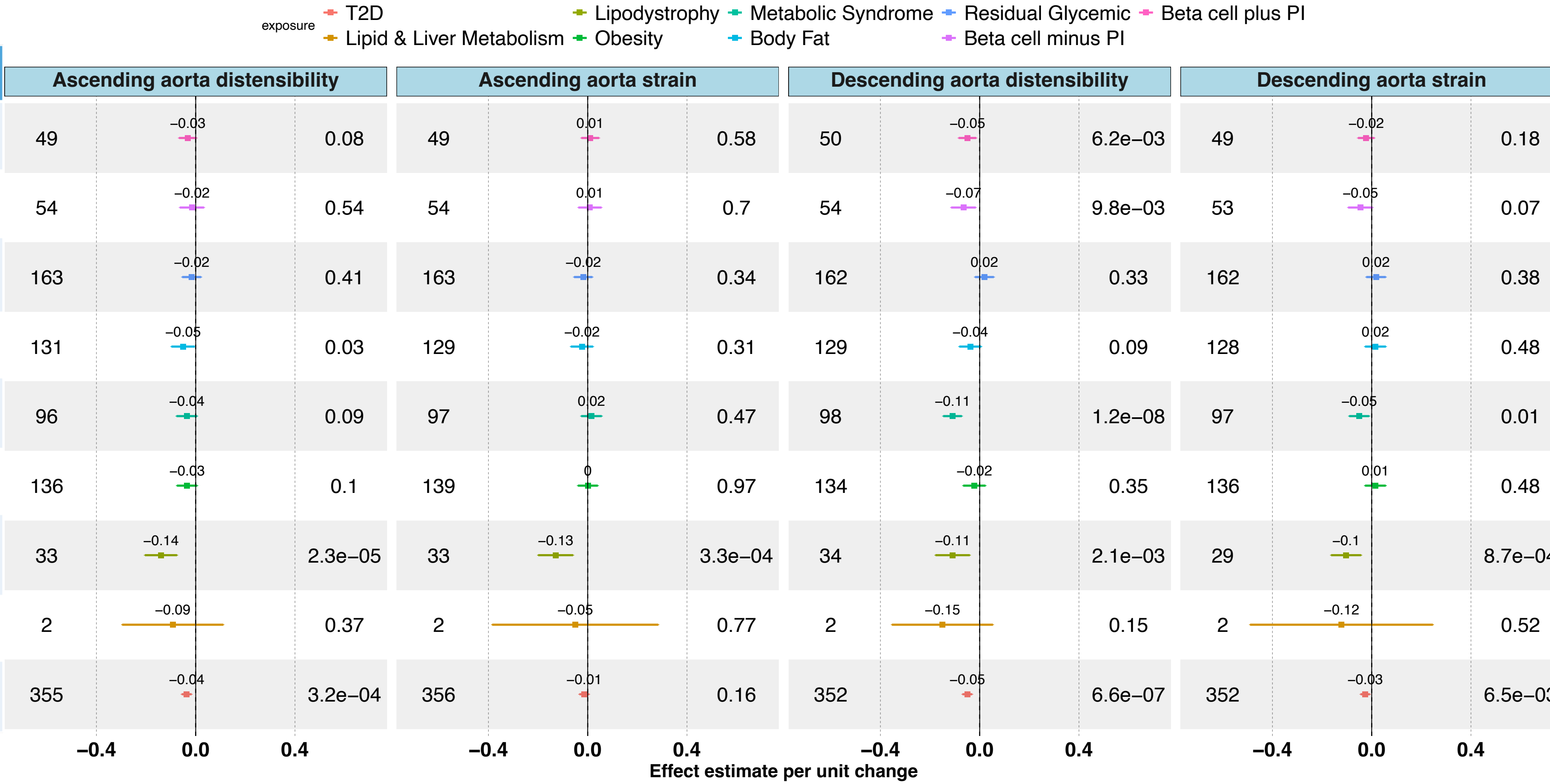


Figure S5: Univariate MR (IVW) for T2D predefined mechanistic clusters)

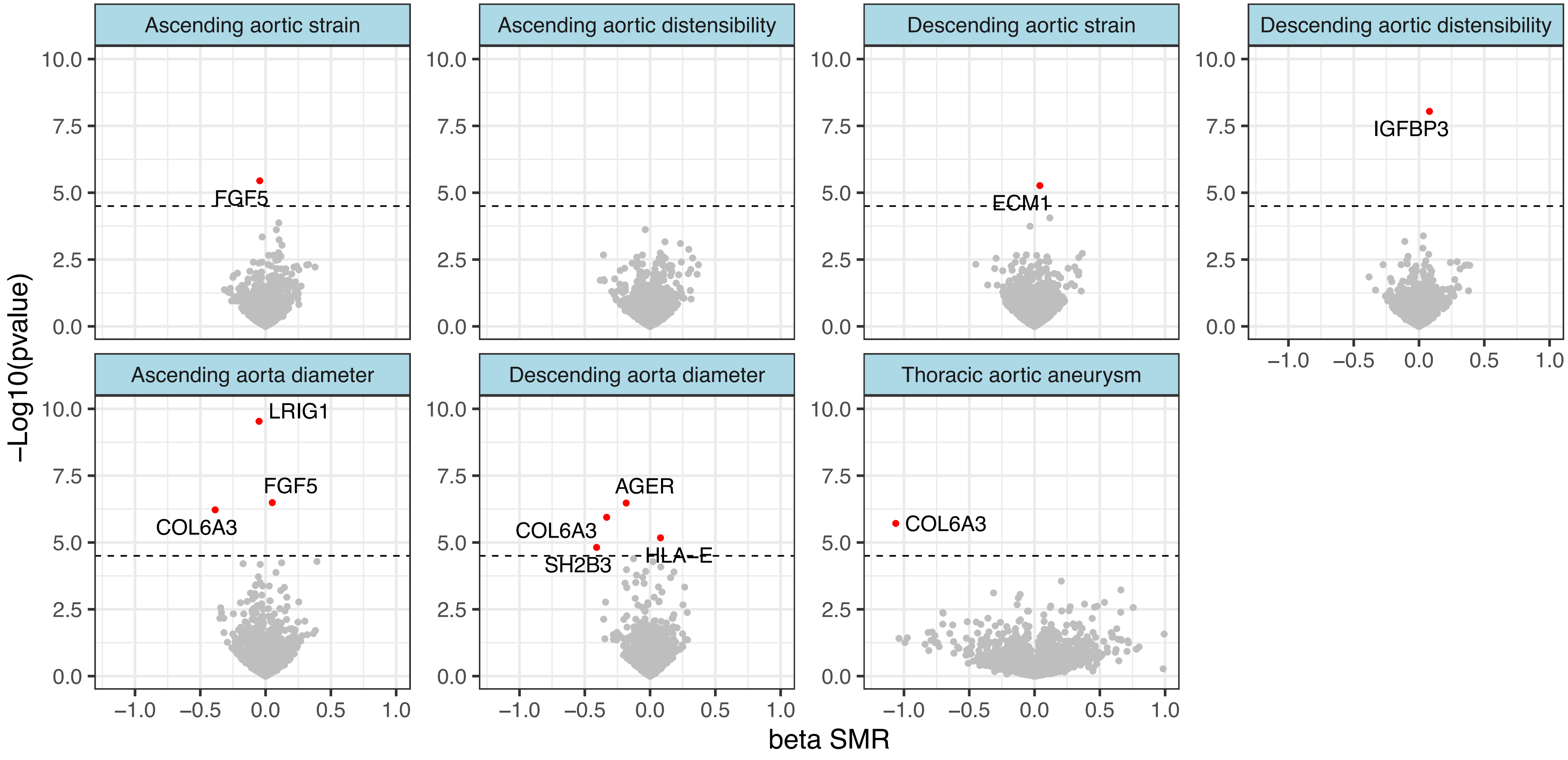


Figure S6: Proteomic MR with aortic phenotypes
